# Agreement between ranking metrics in network meta-analysis: an empirical study

**DOI:** 10.1101/2020.02.11.20021055

**Authors:** Virginia Chiocchia, Adriani Nikolakopoulou, Theodoros Papakonstantinou, Matthias Egger, Georgia Salanti

## Abstract

**Objective:** To empirically explore the level of agreement of the treatment hierarchies from different ranking metrics in network meta-analysis (NMA) and to investigate how network characteristics influence the agreement.

**Design:** Empirical evaluation from re-analysis of network meta-analyses.

**Data:** 232 networks of four or more interventions from randomised controlled trials, published between 1999 and 2015.

**Methods:** We calculated treatment hierarchies from several ranking metrics: relative treatment effects, probability of producing the best value (*p*_*BV*_) and the surface under the cumulative ranking curve (SUCRA). We estimated the level of agreement between the treatment hierarchies using different measures: Kendall’s *τ* and Spearman’s *ρ* correlation; and the Yilmaz *τ*_*AP*_ and Average Overlap, to give more weight to the top of the rankings. Finally, we assessed how the amount of the information present in a network affects the agreement between treatment hierarchies, using the average variance, the relative range of variance, and the total sample size over the number of interventions of a network.

**Results:** Overall, the pairwise agreement was high for all treatment hierarchies obtained by the different ranking metrics. The highest agreement was observed between SUCRA and the relative treatment effect for both correlation and top-weighted measures whose medians were all equal to one. The agreement between rankings decreased for networks with less precise estimates and the hierarchies obtained from *p*_*BV*_ appeared to be the most sensitive to large differences in the variance estimates. However, such large differences were rare.

**Conclusions:** Different ranking metrics address different treatment hierarchy problems, however they produced similar rankings in the published networks. Researchers reporting NMA results can use the ranking metric they prefer, unless there are imprecise estimates or large imbalances in the variance estimates. In this case treatment hierarchies based on both probabilistic and non-probabilistic ranking metrics should be presented.

**STRENGTH AND LIMITATIONS OF THIS STUDY:**

- To our knowledge, this is the first empirical study exploring the level of agreement of the treatment hierarchies from different ranking metrics in network meta-analysis (NMA).
- The study also explores how agreement is influenced by network characteristics.
- More than 200 published NMAs were re-analysed and three different ranking metrics calculated using both frequentist and Bayesian approaches.
- Other potential factors not investigated in this study could influence the agreement between hierarchies.

## INTRODUCTION

Network meta-analysis (NMA) is being increasingly used by policy makers and clinicians to answer one of the key questions in medical decision-making: “what treatment works best for the given condition?” [1,2]. The relative treatment effects, estimated in NMA, can be used to produce ranking metrics: statistical quantities measuring the performance of an intervention on the studied outcomes, thus producing a treatment hierarchy from the most preferable to the least preferable option [3,4].

Despite the importance of treatment hierarchies in evidence-based decision making, various methodological issues related to the ranking metrics have been contested [5–7]. This ongoing methodological debate focuses on the uncertainty and bias in a single ranking metric. Hierarchies produced by different ranking metrics are not expected to agree because ranking metrics differ. For example, a *non-probabilistic ranking metric* such as the treatment effect against a common comparator considers only the mean effect (e.g. the point estimate of the odds-ratio) and ignores the uncertainty with which this is estimated. In contrast, the probability that a treatment achieves a specific rank (a *probabilistic ranking metric*) considers the entire estimated distribution of each treatment effect. However, it is important to understand why and how rankings based on different metrics differ.

There are network characteristics that are expected to influence the agreement of treatment hierarchies from different ranking metrics, such as the precision of the included studies and their distribution across treatment comparisons [4,8]. Larger imbalances in precision in the estimation of the treatment effects affects the agreement of the treatment hierarchies from probabilistic ranking metrics, but it is currently unknown whether in practice these imbalances occur and whether they should inform the choice between different ranking metrics. To our knowledge, no empirical studies have explored the level of agreement of treatment hierarchies obtained from different ranking metrics, or examined the network characteristics likely to influence the level of agreement. Here, we empirically evaluated the level of agreement between ranking metrics and examined how the agreement is affected by network features. The article first describes the methods for the calculation of ranking metrics and of specific measures to assess the agreement and to explore factors that affects it, respectively. Then, a network featuring one of the explored factors is shown as an illustrative example to display differences in treatment hierarchies from different ranking metrics. Finally, we present the results from the empirical evaluation and discuss their implications for researchers undertaking network meta-analysis.

## METHODS

### Data

We re-analysed networks of randomised controlled trials from a database of articles published between 1999 and 2015, including at least 4 treatments; details about the search strategy and inclusion/exclusion criteria can be found in [9,10]. We selected networks reporting arm-level data for binary or continuous outcomes. The database is accessible in the *nmadb* R package [11].

### Re-analysis and calculation of ranking metrics

All networks were re-analysed using the relative treatment effect that the original publication used: odds ratio (OR), risk ratio (RR), standardised mean difference (SMD) or mean difference (MD). We estimated relative effects between treatments using a frequentist random-effects NMA model using the *netmeta* R package [12]. For the networks reporting ORs and SMDs we re-analysed them also using Bayesian models using self-programmed NMA routines in JAGS (https://github.com/esm-ispm-unibe-ch/NMAJags). To obtain probabilistic ranking metrics in a frequentist setting, we used parametric bootstrap by producing 1000 datasets from the estimated relative effects and their variance-covariance matrix. By averaging over the number of simulated relative effects we derived the *probability of treatment i to p roduce t he best value*

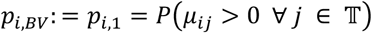

where *μ*_*ij*_ is the estimated mean relative effect of treatment *i* against treatment *j* out of a set 𝕋 of *T* competing treatments. We will refer to this as *p*_*BV*_. This ranking metric indicates how likely a treatment is to produce the largest values for an outcome (or smallest value, if the outcome is harmful). We also calculated the surface under the cumulative ranking curve (*SUCRA*^*F*^) [3]

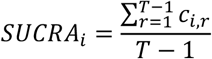

where 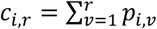 are the cumulative probabilities that treatment *i* will produce an outcome that is among the *r* best values (or that it outperforms *T* − *r* treatments). SUCRA, unlike *p*_*BV*_, also considers the probability of a treatment to produce unfavourable outcome values. Therefore, the treatment with the largest SUCRA value represents the one that outperforms the competing treatments in the network, meaning that overall it produces preferable outcomes compared to the others. We also obtained SUCRAs within a Bayesian framework (*SUCRA*^*B*^).

To obtain the non-probabilistic ranking metric we fitted an NMA model and estimated related treatment effects. To obtain estimates for all treatments we reparametrize the NMA model so that each treatment is compared to a fictional treatment of average performance [13,14]. The estimated relative effects against a fictional treatment *F* of average efficacy 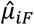 represent the ranking metric and the corresponding hierarchy is obtained simply by ordering the effects from the largest to the smallest (or in ascending order, if the outcome is harmful). The resulting hierarchy is identical to that obtained using relative effects from the conventional NMA model. In the rest of the manuscript, we will refer to this ranking metric simply as relative treatment effect.

### Agreement between ranking metrics

To estimate the level of agreement between the treatment hierarchies obtained using the three chosen ranking methods we employed several correlation and similarity measures. To assess the correlation between ranking metrics we used Kendall’s *τ* [15] and the Spearman’s *ρ* [16]. Both Kendall’s *τ* and Spearman’s *ρ* give the same weight to each item in the ranking. In the context of treatment ranking, the top of the ranking is more important than the bottom. We therefore also used a top-weighted variant of Kendall’s *τ*, Yilmaz *τ*_*AP*_ [17], which is based on a probabilistic interpretation of the average precision measure used in information retrieval [18] (see Appendix).

The measures described so far can only be considered for conjoint rankings, i.e. for lists where each item in one list is also present in the other list. Rankings are *non-conjoint* when a ranking is truncated to a certain *depth k* with such lists called *top-k rankings*. We calculated the Average Overlap [19,20], a top-weighted measure for top-k rankings that considers the cumulative intersection (or *overlap*) between the two lists and averages it over a specified depth (cut-off point) *k* (see Appendix for details). We calculated the Average Overlap between pairs of rankings for networks with at least six treatments (139 networks) for a depth *k* equal to half the number of treatments in the network, *k* = *T/*2 (or ((*T* − 1)) / 2 if *T* is an odd number).

We calculated the four measures described above to assess the pairwise agreement between the three ranking metrics within the frequentist setting and summarised them for each pair of ranking metrics and each agreement measure using the median and the 1^st^ and 3^rd^ quartiles. The hierarchy according to *SUCRA*^*B*^ was compared to that of its frequentist equivalent to check how often the two disagree.

### Influence of network features on the rankings agreement

The main network characteristic considered was the amount of information in the network (reflected in the precision of the estimates). Therefore, for each network we calculated the following measures of information:

- the average variance, calculated as the mean of the variances of the estimated treatment effects *mean*(*SE*^2^), to show how much information is present in a network altogether;
- the relative range of variance, calculated as 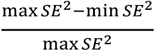, to describe differences in information about each intervention within the same networks;
- the total sample size of a network over the number of interventions.

These measures are presented in scatter plots against the agreement measurements for pairs of ranking metrics.

All the codes for the empirical evaluation are available at https://github.com/esm-ispm-unibe-ch/rankingagreement.

## ILLUSTRATIVE EXAMPLE

To illustrate the impact of the amount of information on the treatment hierarchies from different ranking metrics, we used a network of nine antihypertensive treatments for primary prevention of cardiovascular disease that presents large differences in the precision of the estimates of overall mortality [21]. The network graph and forest plot of relative treatment effects of each treatment versus placebo are presented in Figure 1. The relative treatment effects reported are risk ratios (RR) estimated using a random effects NMA model.

**Figure 1:**
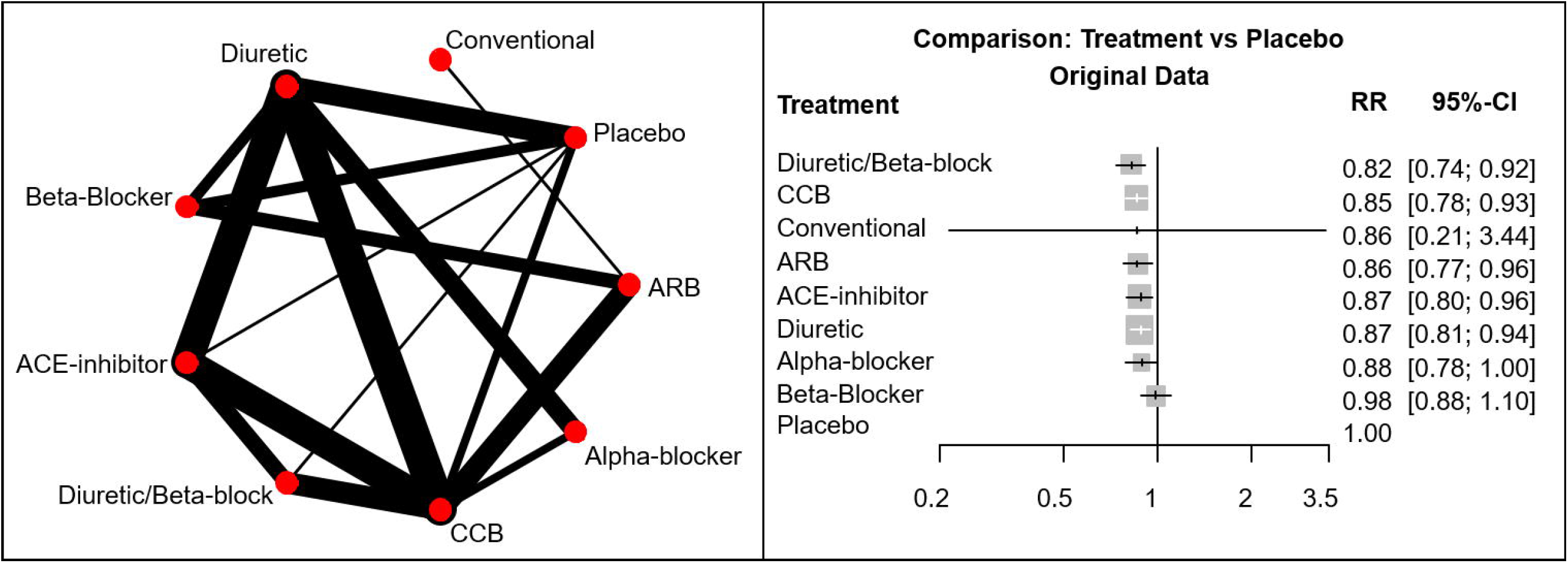
(left panel) Network graph of network of nine antihypertensive treatments for primary prevention of cardiovascular disease [21]. Line width is proportional to inverse standard error of random effects model comparing two treatments. **(right panel) Forest plots of relative treatment effects of overall mortality for each treatment versus placebo**. RR: risk ratio; ACE=Angiotensin Converting Enzyme; CCB=Calcium Channel Blockers; ARB=Angiotensin Receptor Blockers; SE=standard error.

Table 1 shows the treatment hierarchies obtained using the three ranking metrics described above. The highest overall agreement is between hierarchies from the *SUCRA*^*F*^ and the relative treatment effect as shown by both correlation (Spearman’s *ρ* = 0.93, Kendall’s *τ* = 0.87) and top-weighted measures (Yilmaz’s *τ*_*AP*_= 0.87; Average Overlap = 0.85). The level of agreement decreases when *SUCRA*^*F*^ and the relative treatment effect are compared with *p*_*BV*_ rankings (Spearman’s *ρ* = 0.63 and *ρ* = 0.85 respectively). Agreement with *p*_*BV*_ especially decreases when considering top ranks only (Average Overlap is 0.48 for *p*_*BV*_ versus *SUCRA*^*F*^ and 0.54 for *p*_*BV*_ versus relative treatment effect). All agreement measures are presented in online supplementary **Table S1**.

**Table 1:** Example of treatment hierarchies from different ranking metrics for a network of nine antihypertensive treatment for primary prevention of cardiovascular disease [21].

| Treatment | Original data |  |  | Fictional data with increased precision for Conventional treatment versus ARB |  |  |
| --- | --- | --- | --- | --- | --- | --- |
| | $p_{BV}$ ranks | $SUCRA_F$ ranks | Relative treatment effect ranks | $p_{BV}$ ranks | $SUCRA_F$ ranks | Relative treatment effect ranks |
| Conventional | 1 | 6 | 3.5 | 3 | 4 | 3.5 |
| Diuretic/Beta-blocker | 2 | 1 | 1 | 1 | 1 | 1 |
| ARB | 3 | 3 | 3.5 | 4.5 | 3 | 3.5 |
| CCB | 4 | 2 | 2 | 2 | 2 | 2 |
| Alpha-blocker | 5 | 7 | 7 | 4.5 | 7 | 7 |
| ACE-inhibitor | 6 | 4 | 5 | 6.5 | 5 | 5 |
| Diuretic | 7 | 5 | 6 | 6.5 | 6 | 6 |
| Placebo | 8.5 | 9 | 9 | 8.5 | 9 | 9 |
| Beta-Blocker | 8.5 | 8 | 8 | 8.5 | 8 | 8 |

ACE=Angiotensin Converting Enzyme; CCB=Calcium Channel Blockers; ARB=Angiotensin Receptor Blockers. $p_{BV}$ : probability of producing the best value; $SUCRA_F$ : surface under the cumulative ranking curve (calculated in frequentist setting); relative treatment effect stands for the relative treatment effect against fictional treatment of average performance. The first three rankings from the left-hand side are obtained using the original data; the equivalent three rankings on the right-hand side are produced by reducing the standard error of the Conventional versus ARB treatment effect from 0.7 to a fictional value of 0.01.

The reason for this disagreement is explained by the differences in precision in the estimated effects (Figure 1). These RRs versus placebo range from 0.82 (Diuretic/Beta-blocker versus placebo) to 0.98 (Beta-blocker versus placebo). All estimates are fairly precise except for the RR of conventional therapy versus placebo whose 95% confidence interval extends from 0.21 to 3.44. This uncertainty in the estimation is due to the fact that conventional therapy is compared only with Angiotensin Receptor Blockers (ARB) via a single study. This large difference in the precision of the estimation of the treatment effects mostly affects the *p*_*BV*_ ranking, which disagrees the most with both of the other rankings. Consequently, the Conventional therapy is in the first rank in the *p*_*BV*_ hierarchy (because of the large uncertainty) but only features in the third/fourth and sixth rank using the relative treatment effects and *SUCRA*^*F*^ hierarchies, respectively.

To explore how the hierarchies for this network would change in case of increased precision, we reduced the standard error of the Conventional versus ARB treatment effect from the original 0.7 to a fictional value of 0.01 resulting in a confidence interval 0.77 to 0.96. The columns in the right-hand side of Table 1 display the three equivalent rankings after the standard error reduction. The conventional treatment has moved up in the hierarchy according to *SUCRA*^*F*^ and moved down in the one based on *p*_*BV*_, as expected. The treatment hierarchies obtained from the *SUCRA*^*F*^ and the relative treatment effect are now identical (Conventional and ARB share the 3.5 rank because they have the same effect estimate) and the agreement with the *p*_*BV*_ rankings also improved (*p*_*BV*_ versus *SUCRA*^*F*^ Spearman’s *ρ* = 0.89, Average Overlap = 0.85; *p*_*BV*_ versus relative treatment effect Spearman’s *ρ* = 0.91, Average Overlap = 0.94; online supplementary **Table S1**).

## RESULTS

A total of 232 networks were included in our dataset. Their characteristics are shown in Table 2. The majority of networks (133 NMAs, 57.3%) did not report any ranking metrics in the original publication. Among those which used a ranking metric to produce a treatment hierarchy, the probability of being the best was the most popular metric followed by the SUCRA with 35.8% and 6.9% of networks reporting them, respectively.

**Table 2:** Characteristics of the 232 NMAs included in the re-analysis.

| Characteristics of networks | Median | IQR |
| --- | --- | --- |
| Median number of treatments compared | 6 | (5, 9) |
| Median number of studies included | 19 | (12, 34) |
| Median total sample size | 6100 | (2514, 17264) |
|  | Number of NMAs | % |
| Beneficial outcome | 97 | 41.8% |
| Dichotomous outcome | 185 | 79.7% |
| Continuous outcome | 47 | 20.3% |
| Published before 2010 | 42 | 18.1% |
| Ranking metric used in original publication (non-exclusive): |  |  |
| Probability of producing the best value | 83 | 35.8% |
| Rankograms | 7 | 3% |
| Median or mean rank | 3 | 1.3% |
| SUCRA | 16 | 6.9% |
| Other | 2 | 0.9% |
| None | 133 | 57.3% |
| Published in general medicine journals<sup>†</sup> | 125 | 53.9% |
| Published in health services research journals<sup>‡</sup> | 3 | 1.3% |
| Published in specialty journals | 104 | 44.8% |

IQR: interquartile range; NMA: network meta-analysis; SUCRA: surface under the cumulative ranking curve.
<sup>†</sup> Includes the categories Medicine, General & Internal, Pharmacology & Pharmacy, Research & Experimental, Primary Health Care.
<sup>‡</sup> Includes the categories Health Care Sciences & Services, Health Policy & Services.

Table 3 presents the medians and quartiles for each similarity measures. All hierarchies showed a high level of pairwise agreement, although the hierarchies obtained from the *SUCRA*^*F*^ and the relative treatment effect presented the highest values for both unweighted and with top-weighted measures (all measures’ median equals 1). Only 4 networks (less than 2%) had a Spearman’s correlation between *SUCRA*^*F*^ and the relative treatment effect less than 90% (not reported). The correlation becomes less between the *p*_*BV*_ rankings and those obtained from the other two ranking metrics with Spearman’s *ρ* median decreasing to 0.9 and Kendall’s *τ* decreasing to 0.8. The Spearman’s correlation between these rankings was less than 90% in about 50% of the networks (in 116 and 111 networks for *p*_*BV*_ versus *SUCRA*^*F*^ and *p*_*BV*_ versus relative effect, respectively; results not reported). The pairwise agreement between the *p*_*BV*_ rankings and the other rankings also decreased when considering only top ranks (*p*_*BV*_ versus *SUCRA*^*F*^ Yilmaz’s *τ*_*AP*_ = 0.77, Average Overlap = 0.83; *p*_*BV*_ versus relative treatment effect Yilmaz’s *τ*_*AP*_ = 0.79, Average Overlap = 0.88).

The SUCRAs from frequentist and Bayesian settings (*SUCRA*^*F*^ and *SUCRA*^*B*^) were compared in 126 networks (82 networks using the Average Overlap measure) as these reported OR and SMD as original measures. The relevant rankings do not differ much as shown by the median values of the agreement measures all equal to 1 and their narrow interquartile ranges (Table 3). Nevertheless, a few networks showed a much lower agreement between the two SUCRAs. These networks provide posterior effect estimates for which the Normal approximation is not optimal. Such cases were however uncommon as in only 6% of the networks the Spearman’s correlation between *SUCRA*^*F*^ and *SUCRA*^*B*^ was less than 90%. Plots for the Normal distributions from the frequentist setting and the posterior distributions of the log odds-ratios (LOR) for a network with a Spearman’s *ρ* of 0.6 between the two SUCRAs is available in online supplementary **Figure S1** [22].

**Table 3:** Pairwise agreement between treatment hierarchies obtained from the different ranking metrics measured by Spearman ρ, Kendall τ, Yilmaz τ_AP_ and Average Overlap.

| | $p_{BV}$ vs $SUCRA_F$ | $SUCRA_F$ vs relative treatment effect | $p_{BV}$ vs relative treatment effect | $SUCRA_F$ vs $SUCRA_B$ |
| --- | --- | --- | --- | --- |
| **Spearman $\rho$** | 0.9 (0.8, 0.96) | 1 (0.99, 1) | 0.9 (0.8, 0.97) | 1 (0.98, 1) |
| **Kendall $\tau$** | 0.8 (0.67, 0.91) | 1 (0.95, 1) | 0.8 (0.69, 0.91) | 1 (0.93, 1) |
| **Yilmaz $\tau_{AP}$** | 0.78 (0.6, 0.9) | 1 (0.93, 1) | 0.79 (0.65, 0.9) | 1 (0.93, 1) |
| **Average Overlap** | 0.85 (0.72, 0.96) | 1 (0.91, 1) | 0.88 (0.79, 1) | 1 (0.94, 1) |

Medians, 1<sup>st</sup> and 3<sup>rd</sup> quartiles are reported. $p_{BV}$ : probability of producing the best value; $SUCRA_F$ : surface under the cumulative ranking curve (calculated in frequentist setting); $SUCRA_B$ : surface under the cumulative ranking curve (calculated in Bayesian setting); relative treatment effect stands for the relative treatment effect against fictional treatment of average performance.

Figure 2 presents how Spearman’s *ρ* and the Average Overlap vary with the average variance of the relative treatment effect estimates in a network (scatter plots for the Kendall’s *τ* and the Yilmaz’s *τ*_*AP*_ are available in online supplementary **Figure S2**). The treatment hierarchies agree more in networks with more precise estimates (left hand side of the plots).

The association between Spearman’s *ρ* or Average Overlap and the relative range of variance in a network (here transformed to a double logarithm of the inverse values) are displayed in Figure 3. On the right-hand side of each plot we can find networks with smaller differences in the precision of the treatment effect estimates. Treatment hierarchies for these networks show a larger agreement than for those with larger differences in precision. The plots of the impact of the relative range of variance on all measures are available in online supplementary **Figure S3**.

**Figure 2:**
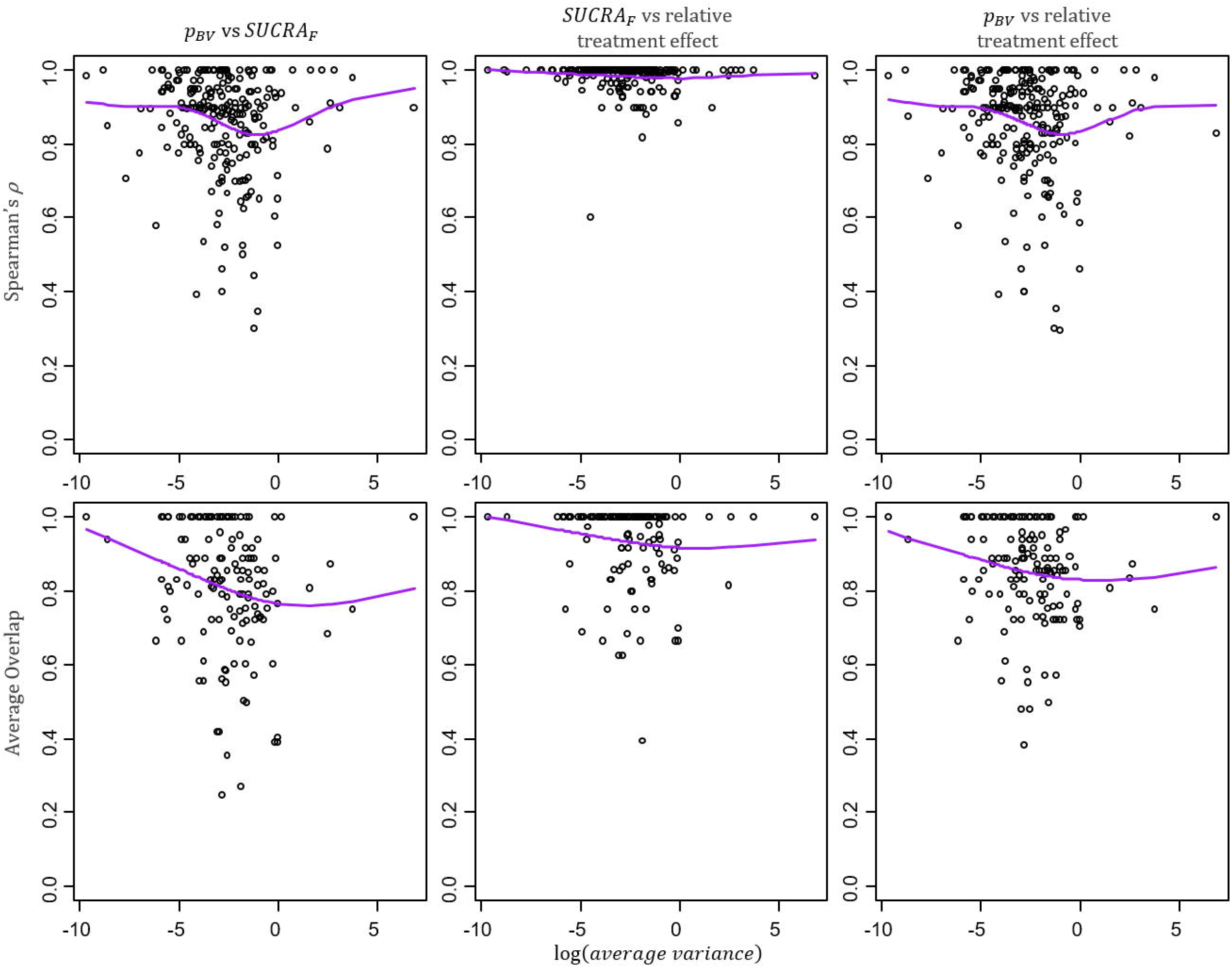
Scatter plots of the average variance in a network and the pairwise agreement between hierarchies from different ranking metrics. The average variance is calculated as the mean of the variances of the estimated treatment effects and describes the average information present in a network. More imprecise network are on the right-hand side of the plots. Spearman ρ (top row) and Average Overlap (bottom row) values for the pairwise agreement between p_BV_ and SUCRA (first column), SUCRA and relative treatment effect (second column), p_BV_ and relative treatment effect (third column). Purple line: cubic smoothing spline with five degrees of freedom.

**Figure 3:**
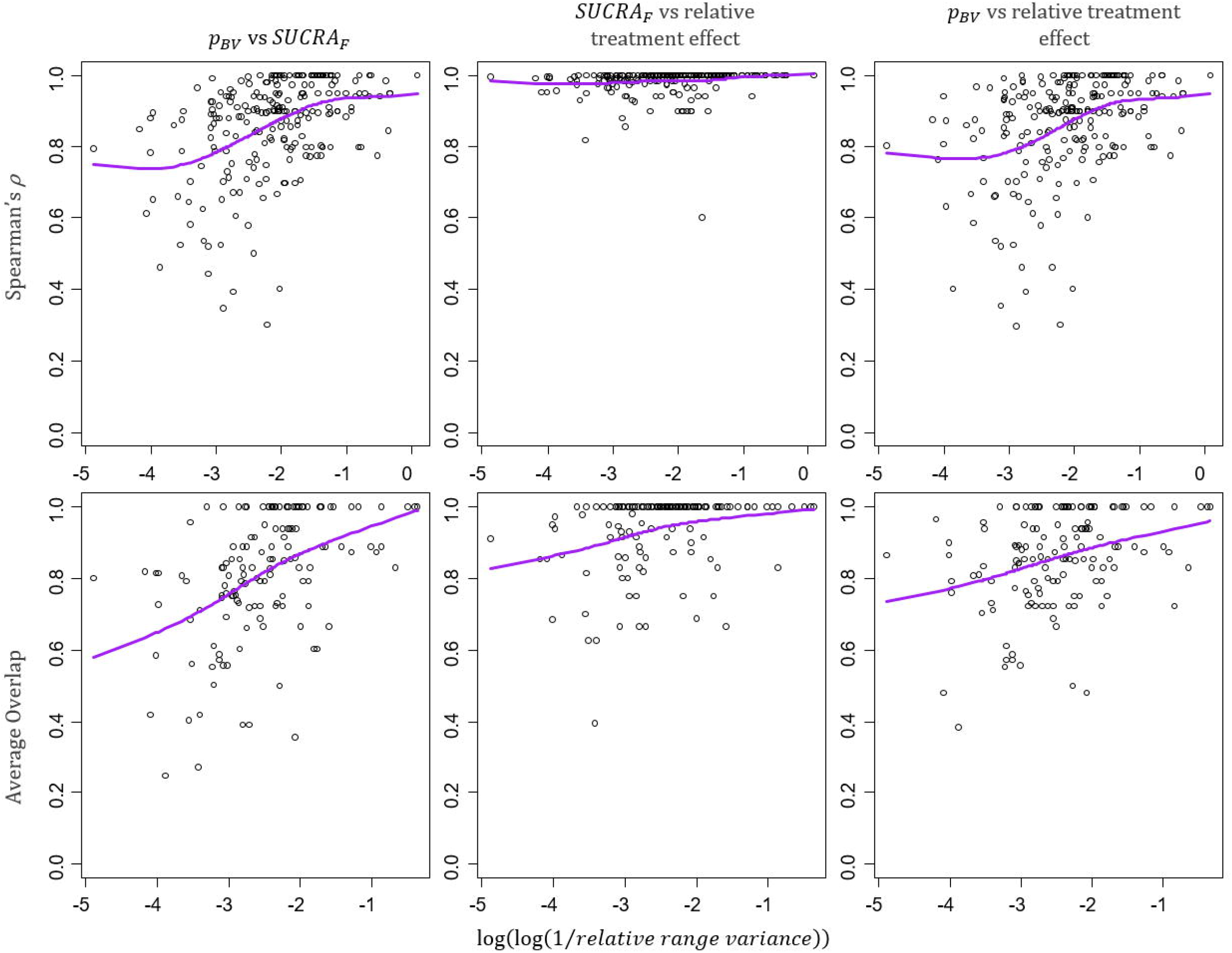
Scatter plots of the relative range of variance in a network and the pairwise agreement between hierarchies from different ranking metrics. The relative range of variance, calculated as 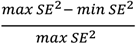, indicates how much the information differs between interventions in the same networks. Networks with larger differences in variance are on the left-hand side of the plots. Spearman ρ (top row) and Average Overlap (bottom row) values for the pairwise agreement between p_BV_ and SUCRA (first column), SUCRA and relative treatment effect (second column), p_BV_ and relative treatment effect (third column). Purple line: cubic smoothing spline with five degrees of freedom.

The total sample size in a network over the number of interventions has a similar impact on the level of agreement between hierarchies. This confirms that the agreement between hierarchies increases for networks with a large total sample size compared to the number of treatments and, more generally, it increases with the amount of information present in a network (online supplementary **Figure S4**).

## DISCUSSION

Our empirical evaluation showed that in practice the level of agreement between treatment hierarchies is overall high for all ranking metrics used. The agreement between treatment hierarchies from *SUCRA* and relative treatment effect was very often perfect. The agreement between the rankings from *SUCRA* or relative treatment effect and the ranking from *p*_*BV*_ was good but decreased when the top-ranked interventions are of interest. The agreement is higher for networks with precise estimates and small imbalances in precision.

Several factors can be responsible for imprecision in the estimation of the relative treatment effects in a network:

- large sampling error, determined by a small sample size, small number of events or a large standard deviation;
- poor connectivity of the network, when only a few links and few closed loops of evidence connect the treatments;
- residual inconsistency;
- heterogeneity in the relative treatment effects.

Random-effects models tend to provide relative treatment effects with similar precision as heterogeneity increases. In contrast, in the absence of heterogeneity when fixed-effects models are used, the precision of the effects can vary a lot according to the amount of data available for each intervention. In the latter case, the ranking metrics are likely to disagree. Our results also confirm that a treatment hierarchy can differ when the uncertainty in the estimation is incorporated into the ranking metric [8,23] and that rankings from the *p*_*BV*_ seem to be the most sensitive to differences in precision in the estimation of treatment effects. We showed graphically that the agreement is less in networks with more uncertainty and with larger imbalances in the variance estimates. However, we also found that such large imbalances do not occur frequently in real data and in the majority of cases the different treatment hierarchies have a relatively high agreement.

We acknowledge that there could be other factors influencing the agreement between hierarchies that we did not explore, such as the risk of bias [23,24] and the chosen effect measures [25]. However, we think it is unlikely that such features play a big role in ranking agreement unless assumptions are violated or data in the network is sparse [26].

To our knowledge, this is the first empirical study assessing the level of agreement between treatment hierarchies from ranking metrics in NMA and it provides further insights into the properties of the different methods. In this context, it is important to stress that neither the objective nor the findings of this empirical evaluation imply that a hierarchy for a particular metric works better or is more accurate than one obtained from another ranking metric. The reason why this sort of comparison cannot be made is that each ranking metric address a specific treatment hierarchy problem. For example, the *SUCRA* ranking addresses the issue of which treatment outperforms most of the competing interventions, while the ranking based on the relative treatment effect gives an answer to the problem of which treatment is associated with the largest average effect for the outcome considered.

Our study shows that, despite theoretical differences between ranking metrics and some extreme examples, they produce very similar treatment hierarchies in published networks. In networks with large amount of data for each treatment, hierarchies based on SUCRA or the relative treatment effect will almost always agree. Large imbalances in the precision of the treatment effect estimates do not occur often enough to motivate a choice between the different ranking metrics. Therefore, our advice to researchers presenting results from NMA is the following: *if the NMA estimated effects are precise*, to use the ranking metric they prefer; *if at least one NMA estimated effect is imprecise*, to refrain from making bold statements about treatment hierarchy and present hierarchies from both probabilistic (e.g. SUCRA or rank probabilities) and non-probabilistic metrics (e.g. relative treatments effects).

## Data Availability

The data for the network meta-analyses included in this study are available in the database accessible using the *nmadb* R package.

## Author contributions

VC designed the study, analysed the data, interpreted the results of the empirical evaluation, and drafted the manuscript. GS designed the study, interpreted the results of the empirical evaluation and revised the manuscript. AN provided input into the study design and the data analysis, interpreted the results of the empirical evaluation and revised the manuscript. TP developed and manages the database where networks’ data was accessed, provided input into the data analysis and revised the manuscript. ME provided input into the study design and revised the manuscript. All the authors approved the final version of the submitted manuscript.

## Patient consent for publication

Not required.

## Data sharing statement

The data for the network meta-analyses included in this study are available in the database accessible using the *nmadb* R package [11].

## APPENDIX

The Yilmaz’s *τ*_*AP*_ calculates the difference between the probability of observing concordance and the probability of observing discordance between two rankings X and Y, penalising more the discordance between top ranks. It can be computed as

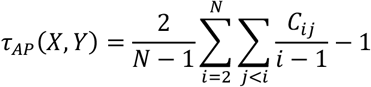

where *c*_*ij*_ is 1 in case the items *i* and *j* are concordant and 0 otherwise; *N* is the total number of items in the ranking.

As Yilmaz’s *τ*_*AP*_ is not symmetric, the authors proposed an alternative measure that takes the average between the two *τ*_*AP*_, with the second being the one calculated after swapping the two rankings

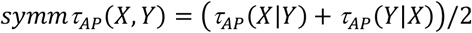

As with the original Kendall’s *τ*, also the Yilmaz’s *τ*_*AP*_ form ula above does not handle ties. Similarly, two formulations to account for this have been proposed [27] and we selected the one that considers correlation as a measure of agreement because more relevant for our purpose. In our chosen version of the Yilmaz’s *τ*_*AP*_, the *τ*_*AP,b*_, neither of the two rankings is considered “true and objective” and ties can be present in either or both of them. The formula appears as follows

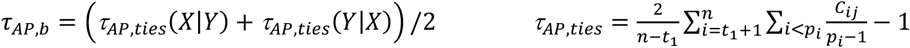

where *t*_*1*_ is the number of items tied in position *i*=1 and *p*_*i*_ is the rank of the first item in *i*’s group.

The Average Overlap is a top-weighted measure for top-k rankings that considers the intersection (or *overlap*) between the two lists, |*X* ∩ *Y*|/*k*. It calculates the cumulative overlap at increasing depths *d, d* ∈ {1…*k*} and average it over the depth (cut-off point) *k*.

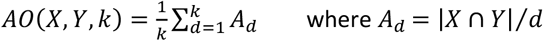

Unlike the previous measures, the average overlap takes values between 0 and 1.

